# High coverage COVID-19 mRNA vaccination rapidly controls SARS-CoV-2 transmission in Long-Term Care Facilities

**DOI:** 10.1101/2021.04.08.21255108

**Authors:** Pablo M De Salazar, Nicholas Link, Karuna Lamarca, Mauricio Santillana

## Abstract

Residents of Long-Term Care Facilities (LTCFs) represent a major share of COVID-19 deaths worldwide. Measuring the vaccine effectiveness among the most vulnerable in these settings is essential to monitor and improve mitigation strategies.

We evaluated the early effect of the administration of BNT162b2 mRNA vaccines to individuals older than 64 years residing in LTCFs in Catalonia, a region of Spain. We monitored all the SARS-CoV-2 documented infections and deaths among LTCFs residents from February 6th to March 28th, 2021, the subsequent time period after which 70% of them were fully vaccinated. We developed a modeling framework based on the relation between community and LTFCs transmission during the pre-vaccination period (July -December 2020) and compared the true observations with the counterfactual model predictions. As a measure of vaccine effectiveness, we computed the total reduction in SARS-CoV-2 documented infections and deaths among residents of LTCFs over time, as well as the reduction on the detected transmission for all the LTCFs.

We estimated that once more than 70% of the LTCFs population were fully vaccinated, 74% (58%-81%, 90% CI) of COVID-19 deaths and 75% (36%-86%, 90% CI) of all expected documented infections among LTCFs residents were prevented. Further, detectable transmission among LTCFs residents was reduced up to 90% (76-93%, 90%CI) relative to that expected given transmission in the community.

Our findings provide evidence that high-coverage vaccination is the most effective intervention to prevent SARS-CoV-2 transmission and death among LTCFs residents. Conditional on key factors such as vaccine roll out, escape and coverage --across age groups--, widespread vaccination could be a feasible avenue to control the COVID-19 pandemic.

## Introduction

Widespread vaccination has the potential to significantly reduce SARS_CoV-2 infections and deaths, and subsequently improve social and economic conditions [1]. Available mRNA COVID-19 vaccines have been approved due to their capacity to reduce symptomatic disease, hospitalizations, and deaths in clinical trials [2,3]; evidence of their real-world effectiveness is growing [4–6], but confirmation still remains limited to certain populations and settings [7–9].

Among all populations, residents of Long-Term Care Facilities (LTCFs) represent a major share of COVID-19 deaths, with a 7-fold higher incidence of death compared to the general population in the US [10,11]. As a consequence, they have been prioritized for vaccinations in most settings. While observational data post-vaccination are still under evaluation at the time of this work, particularly regarding residents of LTCFs [4,12], early assessments on whether clinical trial results are good indicators of vaccine effectiveness in LTCFs would help refine control strategies [1].

In this study, we aimed to quantify the early effect of the administration of the BNT162b2 mRNA COVID-19 vaccine on reducing the risk of SARS-CoV-2 transmission and COVID-19 death among residents of LTCFs in Catalonia (Spain), where high rates of full vaccination among individuals older than 64y (>90% coverage) were reached around 3 months after vaccinations began in December 27, 2020. Prior to the vaccination campaign, the control of SARS-CoV-2 transmission in LTCFs in Catalonia [13] relied on: a) protocolized prevention measures at the individual and facility level, and b) rigorous and timely case ascertainment (including passive case and active case detection) and isolation standards. Protocols regulating the conditions of external visits to the facilities, as well as screening protocols for exits/entries of residents were tightened between December-January [14] but were relaxed once the vaccination campaign was completed. Further, restrictions on individuals’ mobility were implemented by the Catalonian Government at different levels through the territory based on epidemiological risk; in all of Spain, the tightest restrictions were applied in March-June 2020 (which included a country-wide shelter-in-place intervention leading to control of SARS-CoV-2 transmission) and in December 2020-January 2021 [15].

## Methods

The target population analysed in this work was all individuals older than 64y living in care homes in Catalonia, estimated to be around 58,000 in total (see details in supplementary information section S1), between July 2020 and March 2021.This population was vaccinated using the BNT162b2 mRNA COVID-19 vaccine following the guidelines of the Spanish Ministry of Health. The epidemiological data used in this study was collected from --a publicly available repository provided by--the Health Department depending on the Generalitat de Catalunya, the Government of Catalonia. Data on LTCF are collected and updated on a daily basis using health reports from the Primary Care Clinical Station (Care home census), Aggregated Care Health Register (PCR results and deaths), Orfeu, the program for registering virological test results in care homes, and the Catalan Shared Clinical Record where vaccination are registered.

We defined three COVID-19 outcomes to evaluate vaccine efficacy: a) documented infections, comprised of all new infections reported during the study period, independent of symptoms and vaccination status, b) documented deaths, comprised of all deaths attributable to COVID-19 reported during the study period, also independent of vaccination status, and c) detected county-level transmission (herein detected transmission, used as a binary indicator of transmission) defined as at least one documented infection in any facility within a county per unit of time. Each case was assigned the date of diagnostic or testing as the reporting date. LTCFs residents were defined as those living in a LTCF and older than 64y. The general population, or “community”, was defined as all people in a specific area not living in LTCFs. All vaccinated individuals received the two doses of the BNT162b2 mRNA vaccine. Documented COVID-19 infections in LTCFs over time, identified with a molecular test (PCR or antigen test), were assumed to capture most infections given the surveillance protocols in place. Beginning in July 2020, the ascertainment of cases in LTCFs in Catalonia included symptomatic surveillance, tight outbreak investigation and testing of all contacts upon identification of one single case, as well as regular screening of all individuals and staff of a facility independently of symptoms (see further details on supplementary information, Section S1). Documented deaths attributable to COVID-19 included those with laboratory confirmation and those only meeting clinical and epidemiologic criteria. We used 3 spatial resolutions for our analysis, determined by the level of aggregation in the data: a) county level, which corresponds to the definition and boundaries of each “comarca” (n= 41) b) health care area level, which corresponds with the definition and boundaries of each “regió sanitaria” (n=9), and c) regional level, which refers to the largest spatial resolution corresponding to the whole Autonomous Community of Catalonia. For further details see Supplementary Information Section S1.

We generated multiple time series of daily confirmed infections, deaths, and vaccinations in LTCFs, aggregated by healthcare area level, and by the regional (highest aggregation) level. Similarly, we collected daily confirmed infections in the general population, at the healthcare-area level and regional level. For each of these time series, we took a moving weekly average around each day to smooth the daily variation of reporting. Further, we generated a time series of the detected county-level transmission by week. The pre-vaccination period was from July 6 to December 27 2020, when vaccination in LTCFs began. We evaluated the impact of vaccines during an evaluation time period from February 6 to March 28, 2021, the subsequent time period after which 70% of residents were vaccinated with two doses. The 70% threshold was chosen to represent the estimated herd immunity - the estimated level of immunity in a population that prevents uncontrolled spread of infections [16]. As a sensitivity test, we evaluated the effect of partial vaccination in a longer period starting on January 14, when 70% of residents had received the first dose of the vaccine.

We built regression models to predict the number of infections and deaths among LTCFs residents using community infections as inputs. These models were calibrated during the pre-vaccination period and then used to generate predictions in the absence of vaccines during the evaluation period. We compared the models’ counterfactual predictions with observations; discrepancies were used to quantify the effects of the vaccine. Further, for each county with at least one pre-vaccination week with a transmission event in LTCFs and at least one week without one (n=36), we built a logistic regression model to estimate the probability that at least one documented transmission would be observed among LTCFs residents in a given week, using community infections as input. These models were trained on the pre-vaccination period and then used to predict transmission events in the evaluation period among LTCFs residents; the aggregated county-level predicted probabilities and the aggregated observed transmissions were compared to quantify vaccine effectiveness in LTCFs. See Supplementary Information Section S1 for further details on the models.

## Results

We quantified the effect of administering the BNT162b2 vaccine (a) on reducing deaths and documented infections among LTCFs residents older than 64y, and (b) on reducing detected transmission caused by SARS-CoV-2 in LTCFs in Catalonia.

Figure 1 A shows the temporal evolution of infections documented in the community and in LTCFs between July 6, 2020 - March 28 2021. For context, the cumulative vaccination coverage among all LTCFs residents is shown in panel B. Vaccination was deployed among residents and healthcare workers at similar times across LTCFs facilities in the region, beginning December 27 and reaching more than 95% of 2-dose coverage within 2 months.

**Figure 1.**
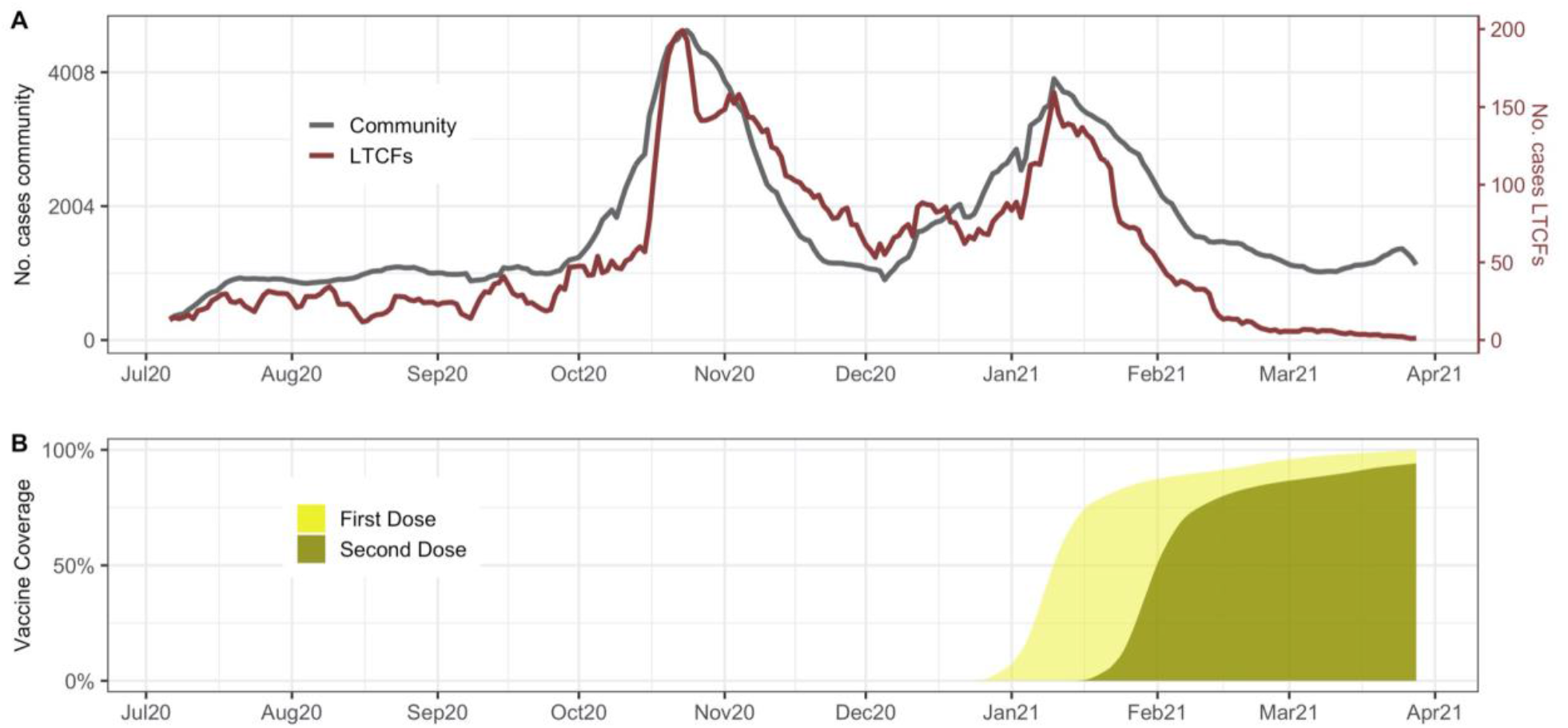
A) Comparison of the total community (grey) and LTCFs documented infections (red) trajectories in Catalonia, Spain. B) First and second dose vaccine coverage among LTCFs residents

Figure 2 A and B show predictions and observations of documented infections and deaths in all of Catalonia over time. We estimated that between February 6-March 28 2021, vaccines prevented 75% of documented infections (36% - 86%, 90% CI), and 74% of deaths (58% - 81%, 90% CI). As well, our analysis shows that two weeks after 70% of residents were fully vaccinated, detected transmission was significantly reduced by 69% (24-80% 90%CI), 54% (0-70%), 50% (0-68%), 69% (25-80%), and 90% (76-93% 90%CI) for each subsequent epidemiological week (Figure 2C).

**Figure 2:**
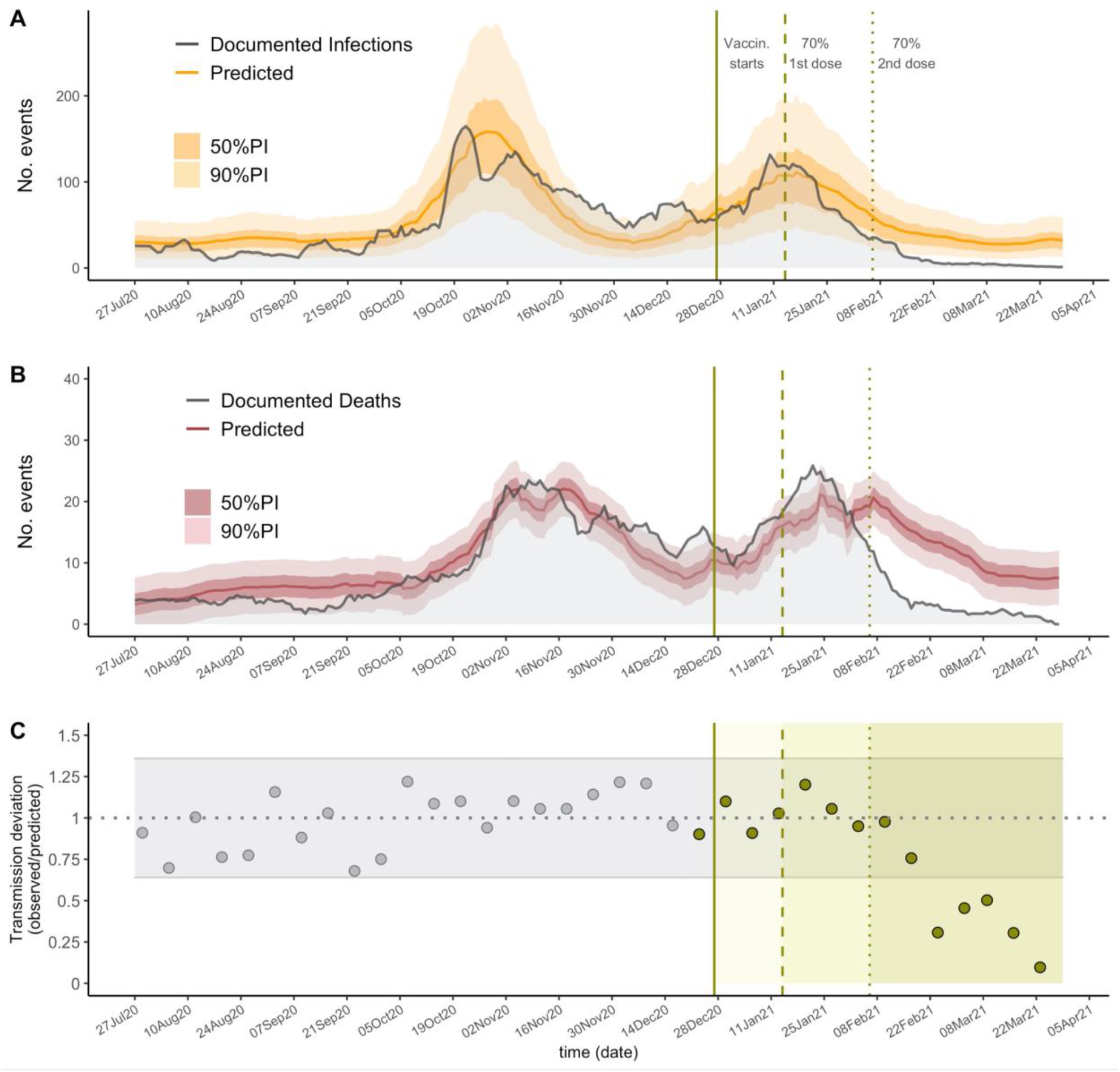
The predictions for infections (A) and deaths (B) across all of Catalonia. The solid lines show the model predictions from training July 6, 2020 through December 27, 2020, the darker shaded background shows the 50% prediction intervals (PI) and the lighter background shows the 90% PI. Vertical lines show key analysis time points: when vaccination started (solid), when 70% of residents received the first dose and when 70% of residents received the second dose. (C) The ratio between observed and predicted transmission at county level in Catalonia, represented by point estimates, grey for the training period and green for the prediction period; grey horizontal ribbon represents the 90% confidence interval. Solid green areas represent the prediction periods after vaccination starts.

## Discussion

In this study we showed that high vaccination coverage (over 70%) prevented around 3 out of 4 expected COVID-19 deaths among residents of LTCFs in subsequent weeks, which is consistent with the vaccine effect on disease severity observed in clinical trials [2] and mortality reduction in other observational studies [6,12]. Further, we found a reduction in transmission after high vaccination coverage was reached while there still was transmission risk (spilled-over from the mainly unvaccinated community transmission), which is caused by both a reduction in vaccinated individuals’ probability of getting infected and a reduction in their probability of transmitting the virus [17].

LTCFs represent enclosed populations, where transmission is caused both by external introduction of the virus, mostly by the staff [18], and internal transmission. Observational findings from facilities with high infection-ascertainment, such as those studied here and elsewhere [4], may provide valuable information of what may be expected to happen in more general settings and populations. Given this, we conclude that beyond the specific risk factors of the population in LTCFs, such as age or comorbidities, high-coverage vaccination can rapidly control SARS-CoV-2 transmission in an enclosed population. Particularly, our analysis suggests that transmission is reduced 3-10-fold one month after vaccination has reached 70% coverage. Of note, our estimates of infections and deaths do not differentiate between infections in vaccinated or unvaccinated individuals, and therefore can be interpreted as the population-level effect of vaccination.

Our methodological assumptions may lead to underestimation of the true vaccine effect. First, our definition of transmission events at the county-level does not capture the facility-level transmission; thus, our results may fail to capture more dramatic facility-level reductions in transmission. Second, the counterfactual estimates of deaths produced by our models may be underestimations of mortality during the evaluation period. Indeed, the documented deaths were generally higher than those estimated by our models in December and January (Figure 2B). It is plausible that factors not considered in the model, such as seasonality [19] and/or the spread of more lethal variants [20] may have increased COVID-19 mortality in recent times. Further, as per guidelines, vaccinations in facilities with ongoing transmission were delayed, which would again lead to underestimation of the vaccine effectiveness. On the other hand, it is possible that relaxing of case ascertainment after the vaccination campaign could lead to overestimation of the effect on documented infections and detected transmission (but unlikely for documented deaths); therefore, we restricted our analysis to the period just after vaccination.

We acknowledge that our investigation has limitations. We assume that the rigorous screening standards in LTCFs in Catalonia led to infection ascertainment close to 100% and also that the ascertainment of community infections does not change dramatically over time. While this may appear unrealistic, public health authorities in Catalonia substantially increased the infection screening efforts in LTCFs before the vaccination campaign [21], beginning in December 2020.

Our model is based on the assumption that the time between disease transmission and case identification is not significantly different between individuals in the community and those in LTCFs. Further, regression models were not designed for accurate infection (or deaths) forecasting and as such, they may not fully capture the epidemiological dynamics, such as changes on COVID-19 restrictions policy over time. However, our efforts were focused on proper inference of the expected epidemic trajectory in the absence of vaccination. This goal is achieved as our models appear to reasonably capture the overall dynamics during the pre-vaccination time periods, even at high spatial granularity (Supplementary Information Figure S1-S2).

Finally, there could be unmeasured confounders, such as behavior or policy changes, not captured by our models, that may have changed the dynamics of transmission between the community and LTCFs over time --this may be the case in the health area Alt Pirineu i Aran prior to the beginning of the vaccination campaign (Supplementary Information Figure S1-S2). Nevertheless, tight restrictions on individuals’ mobility for the whole territory were homogeneously implemented beginning December 2020 and during the period of analysis in this work.

In spite of these limitations, our analyses provide evidence that vaccination may be the most effective intervention in controlling SARS-CoV-2 spread and subsequent risk of death available to date. If our findings continue to be confirmed by future studies, then, conditional on important factors such as vaccine roll out, escape and coverage [22,23], widespread vaccination could be shown to be a feasible avenue to control the COVID-19 pandemic.

## Data Availability

All data was obtained from a publicly available repository https://www.dadescovid.cat.

## Data availability

All data was obtained from a publicly available repository https://www.dadescovid.cat.

## Acknowledgements

We thank Rebecca Kahn, Cristina Colls-Guerra and Antoni Plasencia for their technical advice.

## Author’s contributions

PMD, NL, and MS conceived the study and designed and formulated the primary statistical models. PMD and NL implemented the models, wrote the code and performed all of the analysis. All authors contributed to model design, interpretation of results, writing and editing for the manuscript. MS supervised the study.

## Supplementary Information for

### Section S1. Supplementary Methods

#### Details on ascertainment standards

Starting in July 2020, LTCFs in Catalonia implemented rigorous COVID-19 surveillance that did not solely rely on detection of symptomatic cases [1]. All contacts among staff and residents are screened using molecular test (PCR or antigen test) immediately upon confirmation of an index infection in a facility; further, all staff and residents are regularly screened independently of whether the individuals show symptoms or not; public health guidelines require to screen staff every 2-4 weeks depending on the population size where the LTCFs is located. Thus, the amount of documented infection relies on all infections but over symptomatic infections (which have been estimated to be around ∼60% in LTCFs/skilled nursing homes) [2,3]. For our analysis using this dataset, we assume that the number of documented infections and COVID-19 deaths closely approximates the true number of infections and deaths. For example, in a facility where individuals are screened every 2 weeks [4] on top of having symptomatic surveillance and outbreak investigation standards, reporting no documented infections --when in reality there is transmission--would require the full outbreak to remain undetected (i.e all infections occurring within the same outbreak remain asymptomatic), the outbreak to die off within days or weeks without isolation of exposed individuals, and the infections to show no detectable viral RNA in <14 days after infection (the median duration of detectable SARS-CoV-2 in unvaccinated individuals is estimated to be ∼20 days [5]). Similarly, we assume that the event of not detecting any infection in any of the facilities of a county for a whole week closely approximates the true absence of transmission in the county. Deviations from these assumptions are discussed in the main text.

#### Summarized description and justification of inference models

We use three different models to study three different processes/outcomes: (1) the number of infections/cases in long term care homes, (2) the number of deaths in long term care homes, and (3) the probability of any infection occurring within any long-term care home in a county. For (1), to model the number of infections in LTCFs we use a negative binomial regression, which is standard for modeling case count data. For (2), to model the number of deaths in LTCFs, we use a zero-intercept linear regression, since the relationship of deaths and cases appears to be monotonic and linear -and we assume no covid-19 attributable death would happen in the absence of infections. Model (3) is where we use the logistic regression model to estimate binary “transmission events” at the county level. We defined a transmission event if there was any reported infection in that county for that week (i.e., = 1 if there was at least one new infection and = 0 if there were no new infections). Since the outcome is binary and we wish to generate predictions of the probability of this outcome occurring, a logistic regression approach was chosen. We analyzed this outcome since it is indicative of the efficacy of vaccines reducing SARS-CoV-2 transmission. This is because without a reduction in transmission, we would expect to see a reduction in the total number of infections proportional to the number of people vaccinated, but we wouldn’t expect to see a 100% reduction in infections. Using the binary outcome and the logistic regression model allows us to analyze if there are more county-level 100% reductions in infections than we would expect.

#### Estimating Infections and Deaths Averted By Vaccinations

To estimate the total infections and deaths averted, we aggregate data across all of Catalonia. While it might make sense to use point estimates of infections and deaths averted from individual counties, the more granular model fits are not as good and there is not a clear way to combine the uncertainty from these multiple estimates given their temporal and spatial correlation. We trained a negative binomial model (model 1, eq. 1) to predict the nursing home infections, *N*_*d*_, for day *d* using community infections that day and one week prior, *C*_*d*_ and *C*_*d*−7_ respectively. *N*_*d*_ and *C*_*d*_ are moving weekly averages of documented infections.

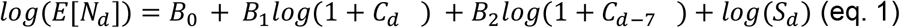

where *S*_*d*_ = 1 − *Im*_*d*_ and *Im*_*d*_ is the proportion of the population that is immune:

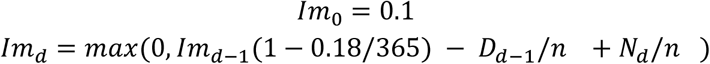

We include an offset term *log*(*S*_*d*_), where *S*_*d*_ is the proportion of the population that is susceptible, to account for the expected decrease in infections due to susceptible depletion, including *log*(*S*_*d*_) amounts to multiplying predictions by the susceptible proportion. *D*_*d*_ is the weekly average of deaths on day *d*, the (1 − 0.18/365) term accounts for a 18% turnover rate in nursing homes in a usual year [6], and *n* is the estimated nursing home population (n=57,922). Since the exact population size in LTCFs was not available at all considered spatial resolutions, we approximated this quantity by the maximum number of vaccinated people once vaccination was reported complete (n=57,922).

For deaths, we use a similar model (model 2, eq. 2), though we use a zero-intercept linear regression of community infections because the relationship between number of infections and number of deaths is likely linear (because a specific proportion of nursing home infections weeks prior will be expected to die).

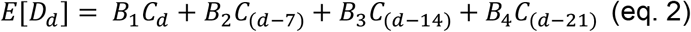

The model was trained on data from a baseline period, July 6, 2020 to December 27, 2020, and then applied to data from an evaluation period, December 28 to March 28 (on December 26, nursing homes began administering vaccines). We generated model fits and prediction intervals in the evaluation period. The prediction intervals for deaths (eq. 2) were generated by using R’s prediction function and the prediction intervals for infections (eq. 1) were generated using a parametric bootstrap procedure. To estimate the number of infections or deaths averted during a target period, we summed the daily model fits and bounds of the prediction intervals. This yields wider intervals than is likely true but given the temporal correlation of the predictions it would not be sensible to combine the variances of each prediction as if they were independent. We selected two target periods to compute averted infections and deaths based on the dates when 70% of the nursing homes residents received the first vaccine dose and when the same proportion received the second dose January 14 and February 6. This allowed us to adjust for changes over time of the vaccines’ effect given the delay between vaccination and efficacious immunization, as well as to account for vaccine coverage close to herd immunity estimates (assumed >70%, see main text). As a supplementary analysis (see supplementary analysis, section S2), we estimated the number of deaths per LTCFs infection, which approximates the mortality rate, before and after vaccination.

#### Predicting change in probability of detected transmission in facilities

We analyzed the changes in detected transmission at the county level. We define the binary outcome of detected transmission occurrence, *O*_*iw*_, as at least one COVID infection among nursing homes residents in county *i* for week *w*. We predict the probability of a transmission occurrence, *π*_*iw*_ = *P*(*O*_*iw*_), using the logistic regression model (model 3, eq.3).

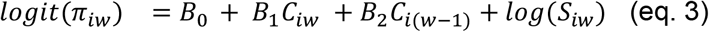

Counties without any detected transmission during the pre-vaccination period (1 county) or without any weeks with no transmission (4 counties) were excluded from this analysis because of the inability to fit a logistic regression model with only one outcome class. To avoid overfitting, we calculated leave-one-out predictions during the pre-vaccination period and out-of-sample predictions for the vaccination period using a model trained in the pre-vaccination period. Then we computed the ratio of observed to predicted transmission events (denoted as *TD*_*w*_ = *Transmission* − *Deviation* (*week w*)) as an approximation of the vaccine effectiveness in fully preventing transmission.

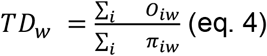

To generate confidence intervals around our predictions, we calculated the sample standard deviation (*σ*_*TD*_) of the pre-vaccination predictions (around a mean ≈1) and use the normal distribution confidence intervals. We believe this is reasonable because we are summing 36 (41 - 5 excluded counties) individual (non-normal) distributions, and by the central limit theorem this should be approximately normal. To estimate the proportion of documented transmission averted with 90% confidence intervals, we calculated 1 − *TD*_*w*_(1 − 1.645*σ*_*TD*_ − *TD*_*w*_, 1 + 1.645*σ*_*TD*_ − *TD*_*w*_).

All analysis was conducted using R 4.0.3 (https://www.R-project.org/).

### Section S2. Predicting change in epidemic size across healthcare areas

Due to the high zero-inflation of documented infections LTCFs at the county level and the difficulty this provides for statistical modeling and inference, we predicted the epidemic size at a higher aggregate level, named “regió sanitaria” (herein healthcare area, n=9). For each area, we predicted LTCFs infections using model 1 and LTCFs deaths using model 2 (see methods). We followed the same procedures as described in the methods to obtain the prediction intervals.

Consistent with the main analysis, healthcare area predictions showed lower-than-expected documented infections and deaths at a more granular level for both target periods of analysis, as seen in eFigure 1 A and B respectively. In 7 out of the 9 health areas the documented infections during the analysis period were consistently lower than the expected infections, and in the two areas where observed infections were higher (Barcelona Ciutat and Metropolitana Nord, among the most heavily populated) they became lower than expected in the final two weeks. Also, infections in Alt Pirineu i Aran become lower than expected around 1-2 weeks earlier than vaccine interventions and remain low for the target time periods, which likely reflect strong specific lockdown measures implemented there. Further, observed deaths were higher during the early analysis period (∼January) in 5 out of 9 healthcare areas (Barcelona ciutat, Camp de Tarragona, Catalunya Central, Girona, Metropolitana Sud), and later became significantly lower, consistent with the main analisis. Factors not considered in the model such as variation of mortality rates due to seasonality or spread of variants with higher lethality might have biased our estimates; in this case, the true number of prevented deaths and infections would be bigger than those estimated by our model. eTable 1 and 2 summarize the number of documented infections and deaths averted by each healthcare area for the two target periods described in the methods. Note that because we used a linear model to predict deaths, some of the predictions are negative, which we truncated at zero. Due to the granularity of the area-level and the greater uncertainty around both models’ predictions, many of the infections- and deaths-averted confidence intervals contain negative values, which would indicate an increase in either outcome. The locations with a greater population (Barcelona Ciutat and Metropolitana Nord) tend to have smaller confidence intervals because their greater population size yields more stable outcome values. While the area-level confidence intervals are wide, when we look at Catalonia as a whole (as in our main analysis) we see a more certain effect of vaccines.

**Figure S1.**
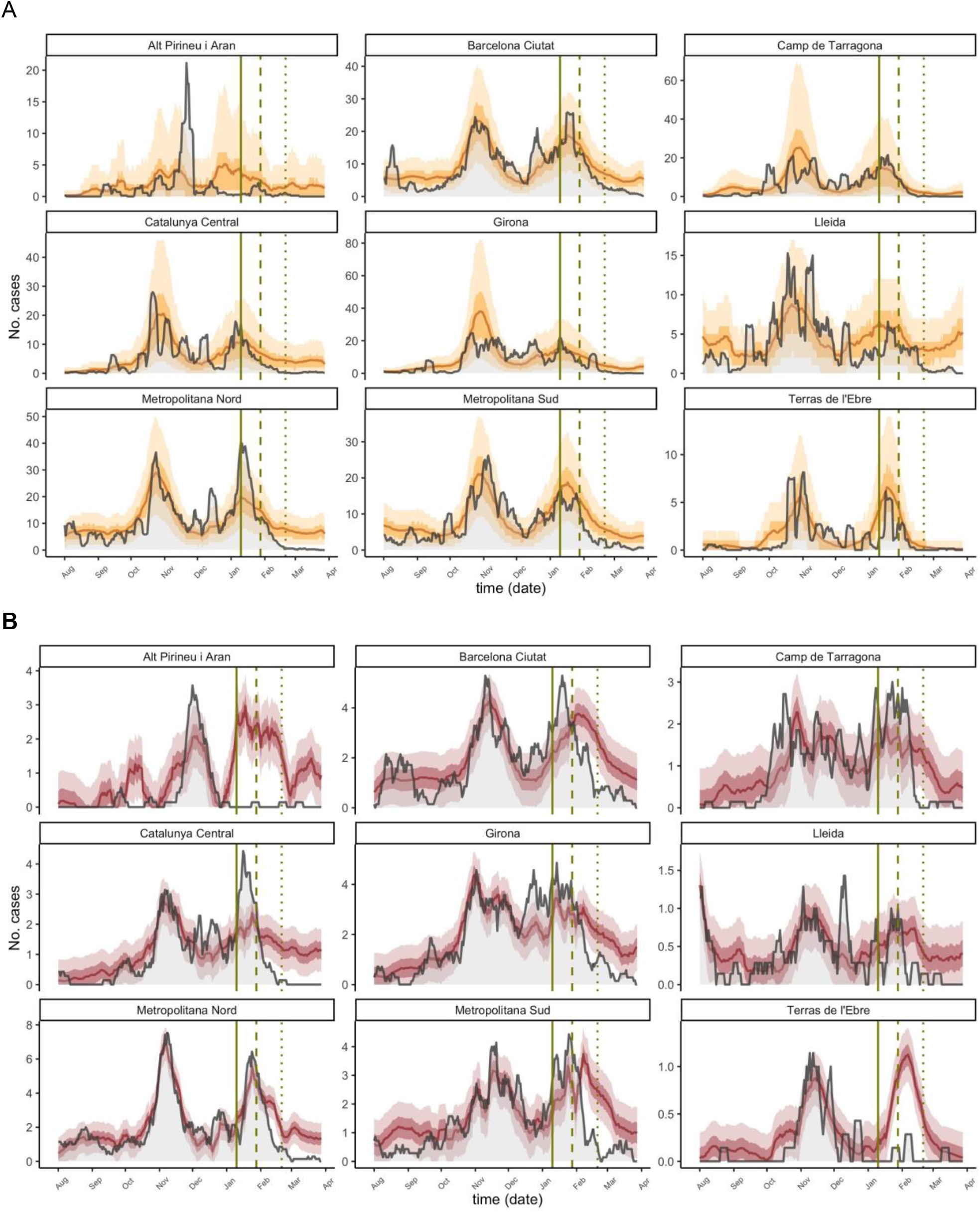
The epidemic size predictions for documented infections by healthcare area. The grey lines are the observed documented infections (A) and deaths (B) and the yellow and red lines are the predicted infections, with ribbons for the IQR and 90% PI. Vertical lines show key analysis time points: when vaccination started (solid), when 70% of residents received the first dose and when 70% of residents received the second dose. Negative predictions are truncated and plotted as 0 for consistency.

**Table S1.** Number of Documented Infections Averted. Number of predicted averted cases in all of Catalon and in each healthcare area. All values are cumulative estimates of cases averted between the starting dates, Jan 14, 2021 and Feb 6, 2021, and March 28, 2021. The 90% confidence intervals are the sums of the bounds of the daily 90% prediction intervals, so they are likely wider than in reality. Negative confidence interval values (indicating higher-than-expected cases) are truncated at 0.

| **Table S1 - Number of Documented Infections Averted** |  |  |
| --- | --- | --- |
|  | **Since 70% first-dose**<br>**vaccination - Jan 14, 2021**<br>**(90% CI)** | **Since 70% second-dose**<br>**vaccination - Feb 6, 2021**<br>**(90% CI)** |
| All of Catalonia<br>*% averted (90% CI)* | 1659 (0, 4817)<br>42% (0%, 68%) | 1371 (157, 2867)<br>75% (36%, 86%) |
| Alt Pirineu i Aran | 109 (0, 469) | 72 (0, 283) |
| Barcelona Ciutat | 174 (0, 753) | 191 (0, 491) |
| Camp de Tarragona | 49 (0, 685) | 87 (0, 304) |
| Catalunya Central | 255 (0, 836) | 195 (0, 527) |
| Girona | 190 (0, 843) | 117 (0, 429) |
| Lleida | 167 (0, 521) | 134 (0, 354) |
| Metropolitana Nord | 256 (0, 854) | 311 (61, 644) |
| Metropolitana Sud | 280 (0, 798) | 182 (0, 437) |
| Terres de l'Ebre | 50 (0, 243) | 11 (0, 69) |

**Table S2.** Number of Deaths Averted. Similar to TableS1, but for deaths averted.

| **Table S2 - Number of Deaths Averted** |  |  |
| --- | --- | --- |
|  | **Since 70% first-dose**<br>**vaccination - Jan 14, 2021**<br>**(90% CI)** | **Since 70% second-dose**<br>**vaccination - Feb 6, 2021**<br>**(90% CI)** |
| All of Catalonia<br>*% averted (90% CI)* | 382 (55, 709)<br>38% (8%, 53%) | 445 (220, 669)<br>74% (58%, 81%) |
| Alt Pirineu i Aran | 117 (49, 185) | 64 (17, 110) |
| Barcelona Ciutat | 56 (0, 135) | 67 (13, 121) |
| Camp de Tarragona | 12 (0, 77) | 31 (-14, 75) |
| Catalunya Central | 37 (0, 89) | 56 (19, 92) |
| Girona | 44 (0, 112) | 58 (11, 105) |
| Lleida | 20 (0, 51) | 18 (-4, 39) |
| Metropolitana Nord | 60 (0, 128) | 71 (24, 119) |
| Metropolitana Sud | 54 (0, 121) | 74 (28, 120) |
| Terres de l'Ebre | 30 (10, 50) | 15 (1, 29) |

### Section S3. Estimating changes in the fatality rates

As sensitivity, we predicted the number of LTCFs deaths from LTCFs documented infections using the model 4 (eq. 5) in Catalonia.

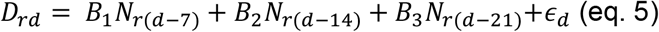

This is a proxy for the fatality rate per infection. Interestingly, the fatality rates seem to increase in January (Figure S2) after vaccines are first delivered and decrease later on, which could explain the excess of deaths observed in the main analysis compared to those predicted from community infections.

**Figure S3.**
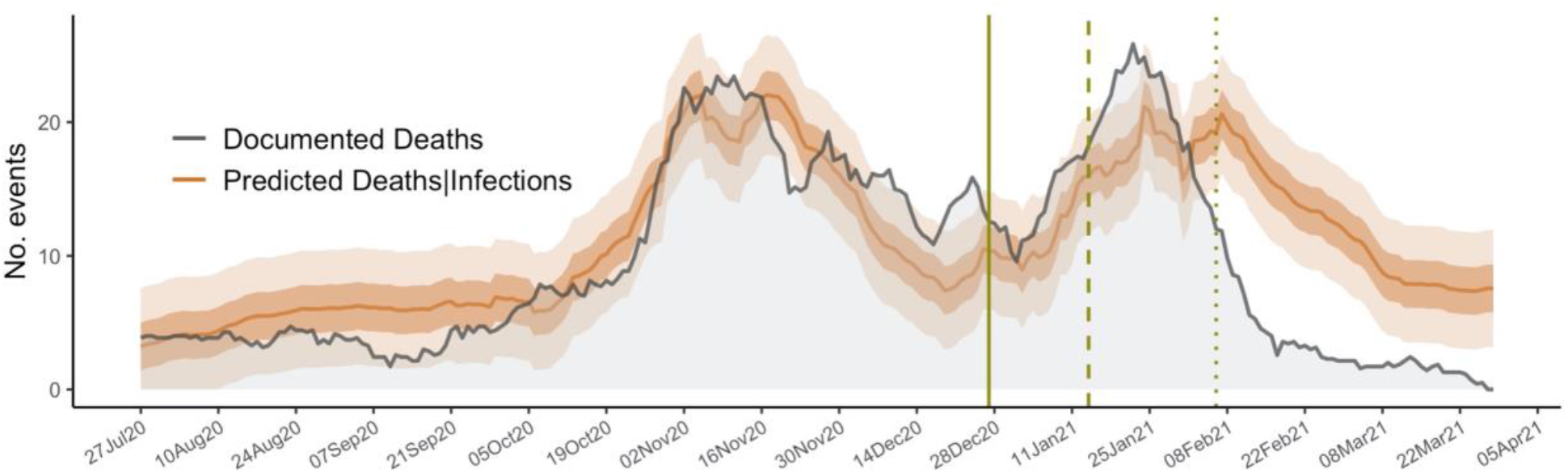
Predictions of the number of LTCFs deaths from LTCFs documented infections using the model 4 (eq. 5) The grey lines are the documented deaths, and the brown lines are the predicted deaths, with ribbons for the 50%PI and 90% PI. Vertical lines show key analysis time points: when vaccination started (solid), when 70% of residents received the first dose and when 70% of residents received the second dose.

### Section S4. Percent Errors for Documented Infections and Death Models

As sensitivity we computed the percent error over time for documented infections (model 1, eq 1) and deaths (model 2, eq 2). As shown in Figure S3 the trend consistently increased toward negative values after vaccination started. For both events, the deviation predictions vs observations during the target time period Feb 6-March 28 becomes close to -100%.

**Figure S4.**
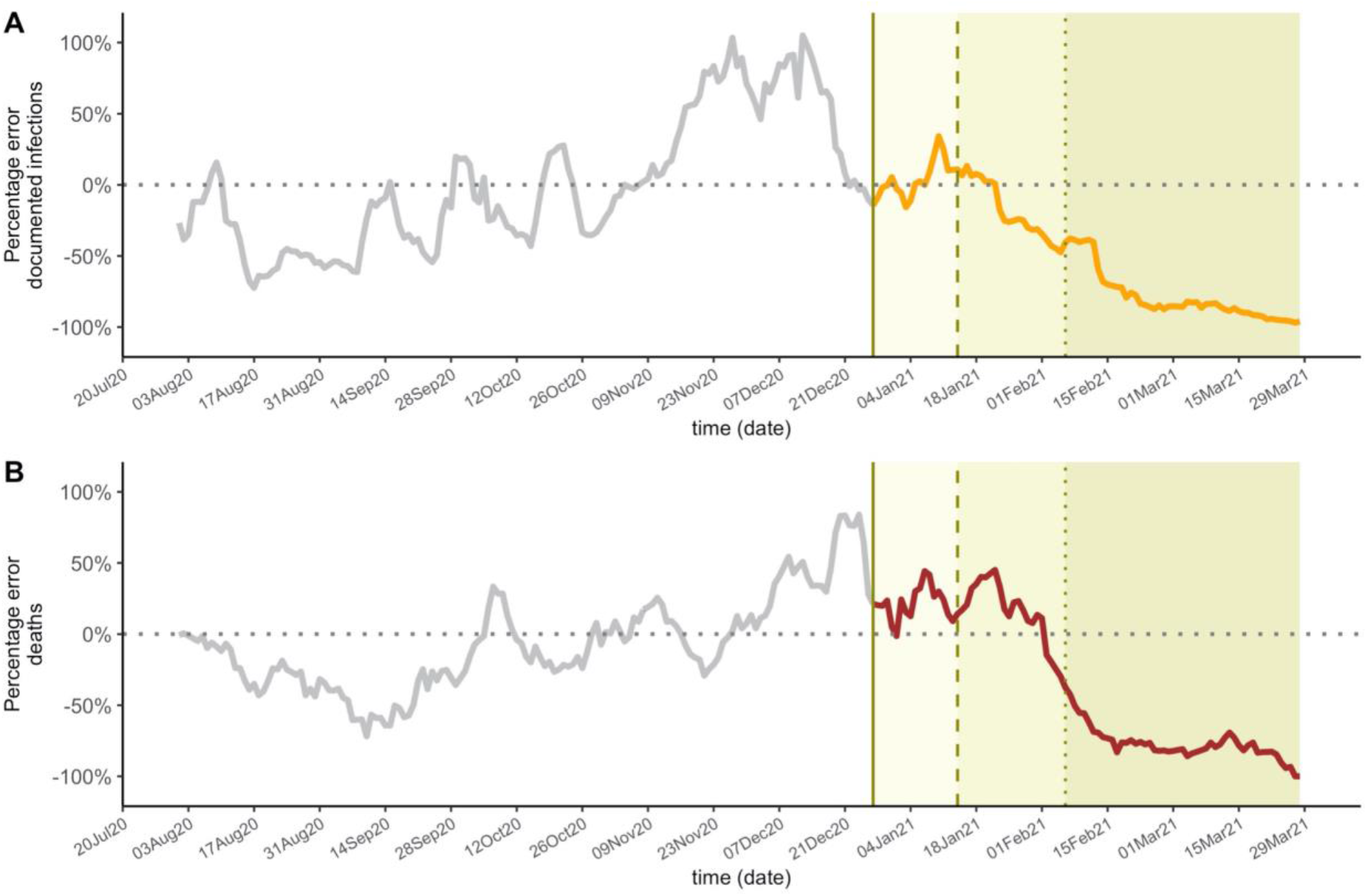
Solid lines represent daily percentage error for documented infections (A) and death model (B). Grey color is used for the training time period pre vaccination and orange and red once vaccination started. Vertical lines show key analysis time points: when vaccination started (solid), when 70% of residents received the first dose and when 70% of residents received the second dose.

